# A biomarker of aging, p16, predicts peripheral neuropathy in women receiving adjuvant taxanes for breast cancer

**DOI:** 10.1101/2022.02.10.22270086

**Authors:** Natalia Mitin, Kirsten A. Nyrop, Susan L. Strum, Anne Knecht, Lisa A Carey, Katherine E. Reeder-Hayes, E. Claire Dees, Trevor A. Jolly, Gretchen G. Kimmick, Meghan S. Karuturi, Raquel E. Reinbolt, JoEllen C. Speca, Erin A. O’Hare, Hyman B. Muss

**Affiliations:** Sapere Bio, Research Triangle Park, NC; School of Medicine, University of North Carolina at Chapel Hill, Chapel Hill, NC; Lineberger Comprehensive Cancer Center, University of North Carolina at Chapel Hill, Chapel Hill, NC; Duke University School of Medicine, Durham NC; MD Anderson Cancer Center, University of Texas, Houston TX; Ohio State University Comprehensive Cancer Center, Columbus OH

## Abstract

**Importance:** Identifying patients at higher risk of chemotherapy-induced peripheral neuropathy (CIPN) is a major unmet need given its high incidence, persistence, and detrimental effect on quality of life.

**Objective:** To determine if expression of p16, a biomarker of aging and cellular senescence, predicts CIPN.

**Design:** Prospective observational cohort study including one hundred fifty-two participants enrolled between January 2014 and August 2018 and followed during the course of adjuvant chemotherapy. Expression of p16 was measured prior to and at the end of chemotherapy. Side effects, including peripheral neuropathy, were assessed prior to each chemotherapy cycle.

**Setting:** A multi-center study including four major academic hospitals and five community oncology clinics.

**Participants:** Women with newly diagnosed with stage I to III breast cancer to receive chemotherapy including a taxane.

**Main Outcomes and Measure:** Development of grade 2+ (moderate or worse) CIPN during the course of chemotherapy. CIPN symptoms were graded by participants’ oncology clinician using the NCI-CTCAE v5 system. Expression of p16 mRNA was measured by qPCR in T-lymphocytes isolated from fresh peripheral blood.

**Results:** A multivariate model including taxane regimen type and p16Age Gap, a measure of discordance between chronological age and p16 expression, identified risk factors for CIPN. Participants with higher chronological age but lower p16 expression prior to chemotherapy (molecularly young) were at the highest risk. Incidence of CIPN positively correlated with chemotherapy-induced increase in p16 expression, with the largest increase seen in participants with the lowest p16 expression prior to treatment.

**Conclusions and Relevance:** This is the first report using a biomarker of senescence in a model to identify patients at risk for taxane-induced CIPN. Studies to confirm and validate our findings are ongoing. When validated, a p16Age Gap-based model can be used to guide chemotherapy selection in patients with early breast cancer and identify patients at high risk who may be candidates for preventive trials.

**Key Points:** *Question:* Is cellular senescence an independent risk factor for chemotherapy-induced peripheral neuropathy?

*Findings:* In a prospective observational cohort study of women with early-stage breast cancer undergoing treatment with taxane chemotherapy, a regression model containing a measure of cellular senescence and taxane type was a statistically significant predictor of grade 2+ CIPN incidence.

*Meaning:* Cellular senescence is an independent risk factor for CIPN that, if validated, could guide treatment selection and identify high-risk patients for preventive strategies.

## INTRODUCTION

Chemotherapy-induced peripheral neuropathy (CIPN) is among the most debilitating, common, and persistent of chemotherapy toxicities^1–3^. CIPN can limit post-treatment quality of life for patients receiving chemotherapy for stages I-III breast cancer^4,5^, since most receive a neurotoxic taxane and will have long-term survival. Up to 30% of these patients experience moderate to severe CIPN^6–8^. Symptoms occur predominantly in the hands and feet and include burning or shooting pain, paresthesia (numbness/tingling), pain perception abnormalities like allodynia and hyper-or hypo-algesia, temperature sensitivity, weakness, and, rarely, ataxia. Multiple studies have shown that moderate to severe CIPN can be dose-limiting, raising concerns about compromised treatment efficacy^7,9,10^.

Even with dose-reductions, over 50% of patients receiving weekly paclitaxel experience limited recovery due to CIPN^11^ with symptoms persisting five or more years post-treatment^3–5^ that dramatically impact quality of life^12–14^. Numbness in the feet can increase the risk of falling, particularly consequential in older patients where fractures can lead to inpatient rehabilitation and loss of independence^4,15–17^. Unfortunately, drug therapies to treat or prevent CIPN are largely ineffective^1,18^, especially among patients receiving taxanes^19^. Patients with severe persistent painful CIPN may also be prescribed opioids, with a risk for opioid addiction^20–22^.

Given the potential severity of symptoms and lack of effective treatments, CIPN prevention becomes critical. Available strategies include cryotherapy^23–26^ and/or selection of a less neurotoxic agent. In Stages I-III breast cancer, neurotoxic taxanes (docetaxel and paclitaxel) are commonly used in both adjuvant and neoadjuvant setting. Paclitaxel confers a much greater risk of CIPN than docetaxel^5,27^, therefore docetaxel may be preferred in at-risk patients. In addition, several preventive pharmacotherapies are under development^28^.

Identifying patients at-risk of CIPN is essential for prevention. Epidemiological risk factors include diabetes, obesity, and age^2,29–31^, though there is no consensus about their relative importance or application in CIPN prevention^2^. Recently, cellular senescence was found to positively associate with cisplatin-induced peripheral neuropathy in mice, and depletion of senescent cells abolished neuropathy^32^.

Cellular senescence is a fundamental mechanism of aging and plays a causative role in nearly all chronic age-related diseases and physical decline^32,33–39^. Senescent cells undergo permanent growth arrest, are resistant to apoptosis, and secrete both inflammatory and pro-fibrotic cytokines, disrupting tissue function and homeostasis^40,41^. Recent studies elegantly demonstrated that induction of senescence in just the immune compartment, and T cells specifically, can induce both senescence and organ damage in tissues throughout the body^42,43^. Expression of p16^INK4a^ (p16) mRNA in peripheral blood T lymphocytes has emerged as a key biomarker of senescence and a measure of senescent cell load^44^. Cellular senescence has not been evaluated as a risk factor for peripheral neuropathy in humans. We hypothesized that p16 expression would associate with the risk of CIPN.

## METHODS

### Study Participants

Women newly diagnosed with stage I-III breast cancer, enrolled in NCT02167932 or NCT02328313, who received a chemotherapy regimen containing a taxane^7^ and had p16 mRNA expression analysis were included in this analysis. Studies were led by the University of North Carolina with REX Healthcare, Ohio State University, MD Andersen, and Duke University participating, and were approved by the IRB of participating sites. The study was performed in agreement with the guidelines of the International Conference on Harmonization, the ethical principles in the Declaration of Helsinki, and all applicable regulations. All patients provided written informed consent before participation in any study-related activities.

### Chemotherapy Regimens and Measures

Chemotherapy regimens were classified as paclitaxel-or docetaxel-containing. CIPN toxicity data was collected as described^7^ For weekly regimens toxicity data was collected every other week so that all toxicity reports were either biweekly or triweekly. Briefly, CIPN symptoms were graded by oncologist using the NCI-CTCAE v5 system. Symptoms were graded as none (0), mild (1), moderate (2), severe (3), or life-threatening (4). Clinicians assessed patients prior to each cycle of chemotherapy.

### p16 expression levels

Peripheral blood samples were collected prior to starting chemotherapy and again at the end of chemotherapy, T cells were isolated, and p16 mRNA expression was analyzed by real-time qPCR as described^45–47^. Positive and negative controls were included in each run; overall precision of p16 measurement (biological and technical) was 0.8 Ct.

### p16Age Gap calculation

P16 expression levels were converted into equivalent years of aging (p16Age) using a linear regression formula derived from analysis of p16 expression in 633 subjects^48^. The p16 value corresponding to the participants’ chronological age at the start of chemotherapy was then subtracted from p16Age to calculate p16Age Gap.

### Statistical Analysis

Chi-square tests, Fisher exact tests, and Student t tests were used to compare patient and clinical characteristics. Statistical significance was set at p = 0.05. Analyses were conducted using SAS/JMP software (SAS Institute Inc, Cary, North Carolina).

To generate a multivariate linear regression model of CIPN risk, chronological age, race, p16 prior to chemotherapy, co-morbidities, and their interactions were tested, as variables may not be independent. Co-morbidities considered were obesity (BMI ≥30), diabetes, peripheral circulatory issues, osteoporosis, arthritis, high blood pressure, emphysema, and liver or kidney disease. Coronary heart disease and stroke were not used due to low prevalence in the CIPN group. Variables and their interactions were considered in the regression model and retained by forward stepwise addition to minimize the Akaike information criterion (AICc). The resulting CIPN probabilities were calculated and plotted. To determine the importance of each variable we calculated indices measuring the importance of factors in a model, in a manner independent of model type and fitting method. The fitted model is used only in calculating predicted values. This method estimates the variability in the predicted response based on a range of variation for each factor. If variation in the factor causes high variability in the response, then that effect is important to the model. Calculations assumed each variable was independent. In this analysis, for each factor, Monte Carlo samples were drawn from a uniform distribution defined by minimum and maximum observed values. Main Effect of importance reflects the relative contribution of that factor alone. Total Effect of importance reflects the relative contribution of that factor both alone and in combination with other factors. To mitigate overfitting when building a neural network model, 1/3 of the data set was reserved as a validation set using holdback function and learning rate of 0.1. The model was built using TanH activation function to fit one hidden layer with 3 nodes. The resulting probabilities of CIPN were calculated and plotted.

For the association between chemotherapy-induced change in p16 and CIPN incidence, p16 expression measured prior to chemotherapy was subtracted from end of treatment expression and stratified into 2 groups: p16 increase (change in p16>0.4, half of assay precision) and no increase (p16 change ≤0.4). Fisher’s exact test (2-taied) was used for group comparison.

## RESULTS

Patient characteristics of 152 study participants with early-stage breast cancer receiving taxane-containing chemotherapies are shown in Table 1. Median age was 56 years (range of 24-83 years), 20% were black, 11% had diabetes, 53% received paclitaxel (48/81 weekly) and the remaining 47% received docetaxel. For analysis purposes, treatments were grouped based on the taxane type, as paclitaxel is more likely to induce CIPN than docetaxel^5,27^. Overall, 29% of participants experienced grade 2 or higher CIPN, of which 82% received paclitaxel and 18% docetaxel-based therapy. In a univariate analysis, none of the patient or clinical characteristics were different between the CIPN and no CIPN groups, including age (p=0.07), diabetes (p=0.7), and p16 expression (p=0.47).

**Table 1.** Demographic and clinical characteristics of the participants in this study.

| Variable | All<br>N=152 | Grade 2+ CIPN<br>n=44 | No CIPN<br>n=108 | P value |
| --- | --- | --- | --- | --- |
| Age - median (SD)<br>Range | 56 (13)<br>24-83 | 59 (11)<br>34-83 | 55 (13)<br>24-77 | 0.07 |
| P16, log2 median (SD) | 9.6 (0.9) | 9.4 (0.9) | 9.5 (0.9) | 0.47 |
| Race, n (%) |  |  |  | 0.19 |
| White | 112 (74) | 32 (73) | 80 (75) |  |
| Black | 30 (20) | 9 (20) | 21 (20) |  |
| Other | 10 (6) | 3 (7) | 7 (5) |  |
| Comorbidities, n (%) |  |  |  |  |
| Diabetes | 15 (11) | 5 (12) | 10 (10) | 0.70 |
| Peripheral vascular issues | 3 (2) | 1 (3) | 2 (2) | 1.00 |
| Osteoporosis | 15 (11) | 3 (7) | 12 (2) | 0.55 |
| Arthritis | 45 (32) | 17 (43) | 28 (28) | 0.09 |
| High blood pressure | 43 (30) | 13 (32) | 30 (30) | 0.84 |
| Coronary heart disease | 5 (4) | 0 (0) | 5 (5) | 0.32 |
| Stroke | 5 (4) | 2 (5) | 3 (3) | 0.63 |
| Emphysema | 4 (4) | 0 (0) | 4 (4) | 0.32 |
| Liver or kidney disease | 10 (7) | 3 (8) | 7 (7) | 1.00 |
| BMI- median (SD) | 28.9 (6.3) | 30.1 (6.6) | 28.5 (6.2) | 0.15 |
| BMI $\geq 30$, n (%) | 59 (39) | 19 (43) | 40 (37) | 0.48 |
| Breast cancer stage, n (%) |  |  |  | 0.15 |
| I | 32 (21) | 5 (11) | 27 (25) |  |
| II | 73 (48) | 25 (57) | 48 (44) |  |
| III | 44 (29) | 13 (30) | 31 (29) |  |
| Missing data | 3 (2) | 1 (2) | 2 (1) |  |
| Hormone receptor and HER-2<br>status, n (%) |  |  |  | 0.86 |
| ER+ or PR+/ HER2- | 71 (47) | 21 (48) | 50 (46) |  |
| HER2+ | 35 (23) | 10 (23) | 25 (23) |  |
| ER-/PR-/HER2- | 46 (30) | 13 (30) | 33 (31) |  |
| Breast cancer surgery, n (%) |  |  |  | 0.94 |
| Lumpectomy | 68 (45) | 19 (43) | 49 (45) |  |
| Mastectomy | 82 (54) | 25 (57) | 57 (53) |  |
| None | 2 (1) | 0 (0) | 2 (2) |  |
| Adjuvant radiotherapy, n (%) |  |  |  |  |
| Yes | 105 (72) | 30 (68) | 75 (74) | 0.55 |
| Chemotherapy regimen, n (%) |  |  |  | 0.0001 |
| Paclitaxel-containing | 81 (53) | 36 (82) | 45 (42) |  |
| Weekly paclitaxel | 48 (32) | 23 (52) | 25 (23) |  |
| Docetaxel-containing | 71 (47) | 8 (18) | 63 (58) |  |
| Regimen<sup>1</sup> |  |  |  |  |
| AC | 52 (34) | 23 (52) | 29 (27) |  |
| AC-TC | 17 (11) | 5 (11) | 12 (11) |  |
| TC | 41 (27) | 2 (5) | 39 (36) |  |
| TC-H | 25 (16) | 5 (11) | 20 (19) |  |
| Other | 17 (11) | 9 (20) | 8 (7) |  |
| Anti-HER2 therapy, n (%) |  |  |  |  |
| Yes | 35 (23) | 10 (23) | 25 (23) | 1.00 |
| Chemotherapy timing, n (%) |  |  |  | 0.59 |
| Neoadjuvant | 64 (42) | 20 (45) | 44 (41) |  |
| Adjuvant | 88 (58) | 24 (55) | 64 (59) |  |
| CIPN-CTCAE, n (%) |  |  |  |  |
| Grade 0 | 53 (35) | 53 (35) | 0 |  |
| Grade 1 | 55 (36) | 55 (36) |  |  |
| Grade 2 | 43 (28) | 43 (28) |  |  |
| Grade 3 | 1 (1) | 1 (1) |  |  |

<sup>1</sup> AC-T- doxorubicin, cyclophosphamide, paclitaxel; AC-TC- doxorubicin, cyclophosphamide, paclitaxel and carboplatin; TC- docetaxel and cyclophosphamide; TC-H docetaxel, carboplatin, and anti-HER2.

Since no variables in Table 1 were independent predictors of CIPN, we tested them in a multivariate regression analysis. Chronological age, race, and co-morbidities were considered. We also included pairwise interactions, as variables like age, p16 and co-morbidities may not be independent (see Methods for details). Taxane type (paclitaxel vs docetaxel) was also included given the difference in CIPN incidence between these two agents. The optimal model from these variables (Model 1) is shown in Table 2. Taxane type, p16 expression before chemotherapy, chronological age, arthritis, and osteoporosis, contributed to model performance.

**Table 2.** Performance and optimal variable combinations produced by multivariate regression analysis to predict risk of grade 2+ CIPN.

|  | Estimate | p | OR | Variable Importance |  |
| --- | --- | --- | --- | --- | --- |
|  |  |  |  | Main Effect | Total Effect |
| Model 1 Variables |  |  |  |  |  |
| Taxane type [paclitaxel] | 0.98 | <0.0001 |  |  |  |
| p16 pre-chemotherapy | -0.67 | 0.02 |  |  |  |
| Arthritis [yes] | 0.62 | 0.03 |  |  |  |
| Age | -0.46 | 0.16 |  |  |  |
| Age*Osteoporosis [yes] | -0.49 | 0.14 |  |  |  |
| Osteoporosis [yes] | 4.83 | 0.16 |  |  |  |
| Age*Arthritis [yes] | -0.03 | 0.23 |  |  |  |
| Model 1 Performance |  |  |  |  |  |
| AICc | 150 |  |  |  |  |
| BIC | 172 |  |  |  |  |
| Chi-square | 34.9 |  |  |  |  |
| p value | 0.0001 |  |  |  |  |
| Model 2 Variables |  |  |  |  |  |
| Taxane type [paclitaxel] | 0.98 | <0.0001 | 7.2 (2.9-17.5) | NA* | NA* |
| p16Age Gap | -0.047 | 0.01 | 0.95 (0.92-0.99) | 0.406 | 0.519 |
| p16 pre-chemotherapy | 1.22 | 0.04 | 3.4 (1.1-10.8) | 0.363 | 0.477 |
| Model 2 Performance |  |  |  |  |  |
| AICc | 161 |  |  |  |  |
| BIC | 173 |  |  |  |  |
| Chi-square | 29.3 |  |  |  |  |
| p value | 0.0001 |  |  |  |  |

Variables tested in Model 1- taxane type, race, BMI ≥ 30, diabetes, peripheral circulatory issues, osteoporosis, arthritis, high blood pressure, emphysema, liver or kidney disease, age, interaction of each co-morbidity and age, and p16 prior to chemotherapy
Variables tested in Model 2- taxane type, race, BMI ≥ 30, diabetes, peripheral circulatory issues, osteoporosis, arthritis, high blood pressure, emphysema, liver or kidney disease, age, interaction of each co-morbidity and age, p16 prior to chemotherapy, and p16Age Gap
\* Variable importance for categorical variables is not calculated

Differences in the behavior of p16 and age in the univariate vs. multivariate analyses prompted us to develop a measure of the difference between p16 measured in the participant prior to treatment and the average p16 level for a non-cancer patient of the same age, called p16Age Gap. First, we developed a method to convert p16 expression from log2 arbitrary units into years, and then directly compared p16-based age and chronological age (see Methods). A distribution of log2 p16, with p16 converted to years (p16Age), and the difference between p16 and chronological age (p16Age Gap) is shown in Figure 1A,B,D. p16Age Gap can also be thought of as a residual in the p16/chronological age regression model. A negative p16Age Gap suggests that an individual has p16 expression levels below an age-appropriate population mean; a p16Age Gap around zero suggests that p16 expression is similar to the population mean; and positive p16Age Gap signifies p16 expression above the population mean.

**Figure 1.**
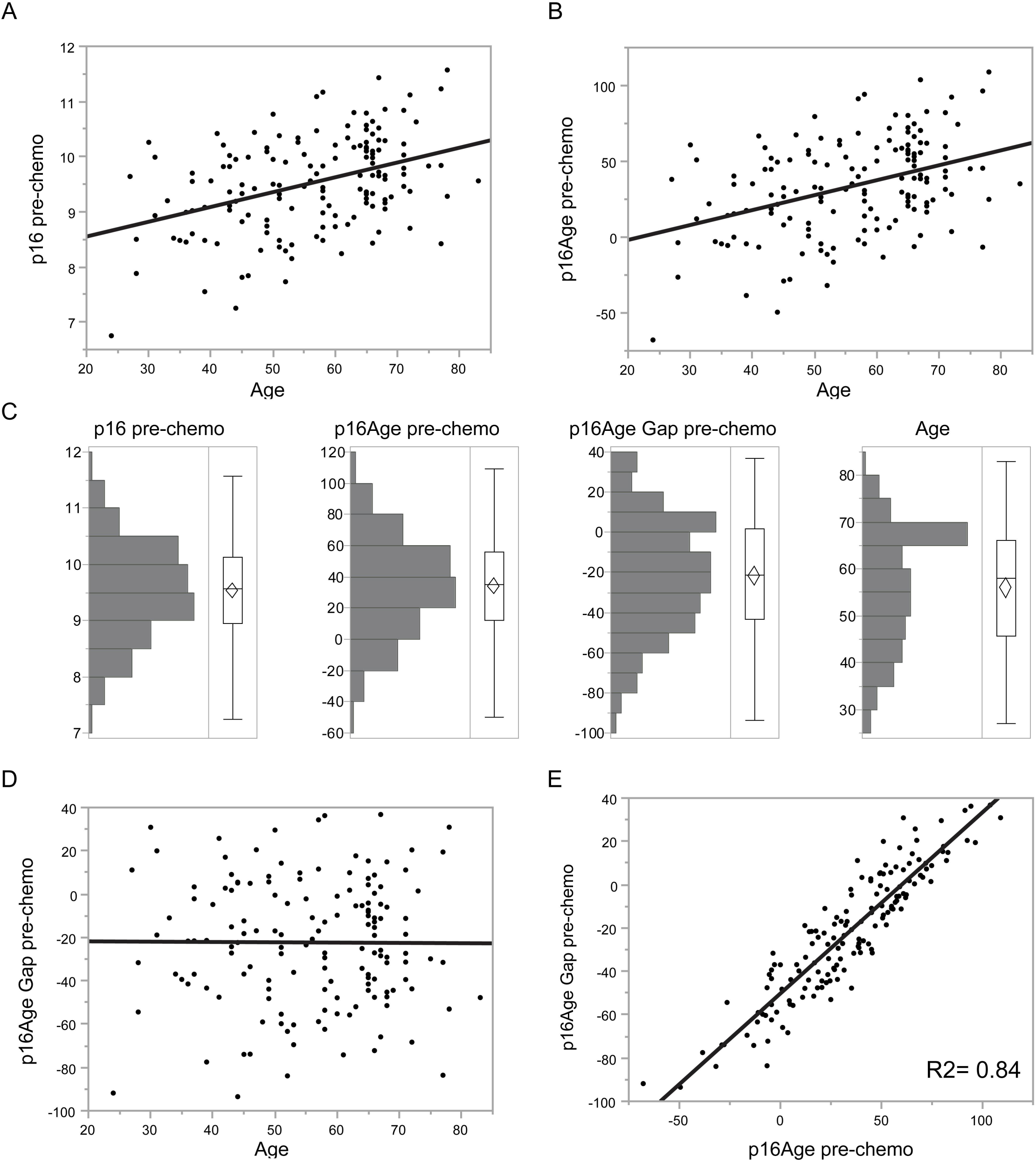
Expession of p16 mRNA, p16Age, and p16Age Gap prior to chemotherapy. Correlation between chronological age and expression levels of p16 mRNA (log2) (A), calculated p16Age (B) and p16Age Gap (D). Distribution of p16, p16Age, p16Age Gap and chronological age is shown in panel C. p16Age (and therefore log2p16) is highly correlated with p16Age Gap.

Distributions of p16 (log2), p16Age, p16Age Gap, and chronological age in this study cohort are shown in Figure 1C and are all normally distributed. Unlike p16Age, p16Age Gap does not correlate with chronological age (p=0.94) (Fig. 1D), demonstrating that participants of all ages may exhibit age-inappropriate levels of p16. Larger absolute values of p16Age Gap values can represent younger participants who are molecularly older (positive p16Age Gap), or older participants who are molecularly younger (negative p16Age Gap). p16Age Gap is strongly associated with p16Age (R2=0.84; p<0.0001; Figure 1E), suggesting that variability in p16 expression, and not differences in chronological age, is the most important contributor to p16Age Gap.

To assess the role of this new measure, p16Age Gap, as a predictor of CIPN risk, we built a second regression model (Table 2, Model 2) using p16Age Gap and the same variables tested in Model 1. The addition of p16Age Gap to these variables, which were used in building Model 1, reduced the number of variables to three, p16Age Gap, p16, and taxane with similar performance to Model 1 (see AICc and BIC). When variables were analyzed for their individual contributions to the CIPN outcome (see Methods), p16Age Gap contributed 41% to the model as an individual predictor and 52% when considered with pre-chemotherapy p16 expression. Patients with a negative p16Age Gap (chronologically older with lower p16 expression) were at a higher risk for CIPN (OR 0.95, p=0.01). Interestingly however, higher pre-chemotherapy p16 was also associated with a higher risk of CIPN in a multivariate model (OR 3.4, p=0.04).

The probability of CIPN in patients receiving paclitaxel-versus docetaxel-based chemotherapy was derived from regression Model 2 and shown in Figure 2. Patients who received paclitaxel had mean probability of grade 2 or greater CIPN of 45% (CI 31-57%) (Figure 2A). Patients who received docetaxel had a mean probability of grade 2 or greater CIPN of 11% (CI 5-17%) (Figure 2B). Correlation between p16Age Gap and probability of CIPN for each taxane is shown in Figure 2C. In patients receiving paclitaxel, the probability of CIPN increased from 14% for patients with p16Age Gap in lower quartile to 48% for patients in the upper quartile.

**Figure 2.**
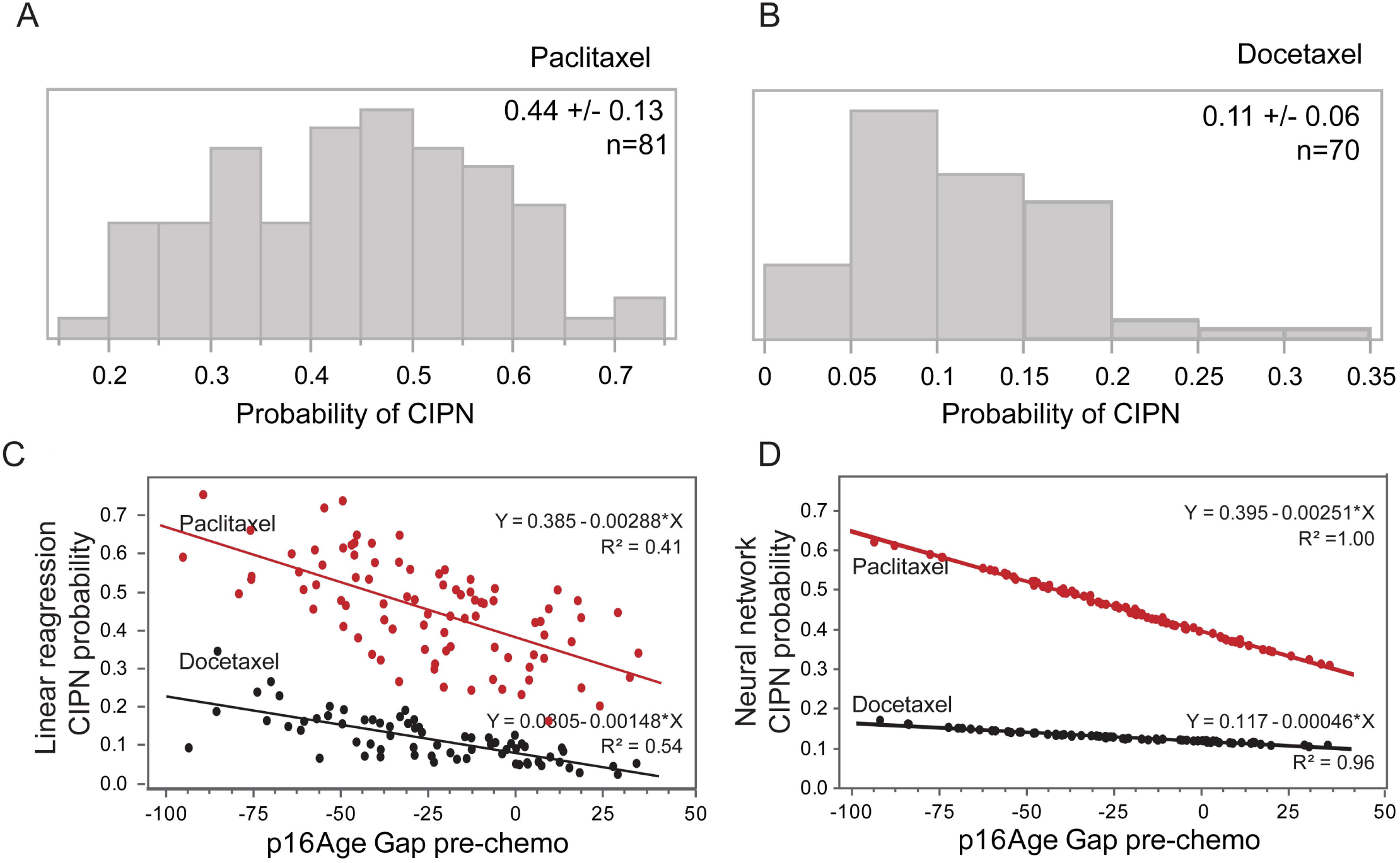
Correlation between p16Age Gap and probability of CIPN. Probability of developing grade 2+ CIPN, calculated from the linear regression Model 2 in participants receiving paclitaxel (A) or docetaxel (B) chemotherapy. Correlation between p16Age Gap single variable and a probability of CIPN derived from linear regression (C) or neural network (D) based model demonstrates significant contribution of p16Age Gap alone to the model.

In addition to the regression model, we used the same variables to build a neural network to predict CIPN (see Methods). The relationship between the probability of CIPN and p16Age Gap as defined by the neural network algorithm is shown in Figure 2D. Interestingly, while the neural network algorithm fit data better than linear regression analysis and produced a much higher R2 value, the slope coefficient for change in CIPN risk for paclitaxel patients was essentially identical between models (linear regression 0.00288, neural network 0.00251). This suggests that the linear regression model was sufficient to describe the relationship between p16Age Gap and CIPN incidence. Taken together, these findings suggest that senescence is an important risk factor for CIPN.

Finally, we asked why patients with lower baseline p16 expression may be at higher risk of CIPN. We previously showed that p16 expression prior to chemotherapy is inversely correlated with the magnitude of chemotherapy-induced p16 increase^49^ (also Figure 3A). Patients who experienced a post-chemotherapy increase in p16 were twice as likely to develop CIPN (37.5% vs 18.3%, p=0.02) as patients whose p16 did not change (Figure 3B). Taken together, our results suggest that participants with lower p16 expression prior to chemotherapy are more likely to have a larger chemotherapy-induced increase in p16 expression and, more importantly, are more likely to experience grade 2 or higher CIPN.

**Figure 3.**
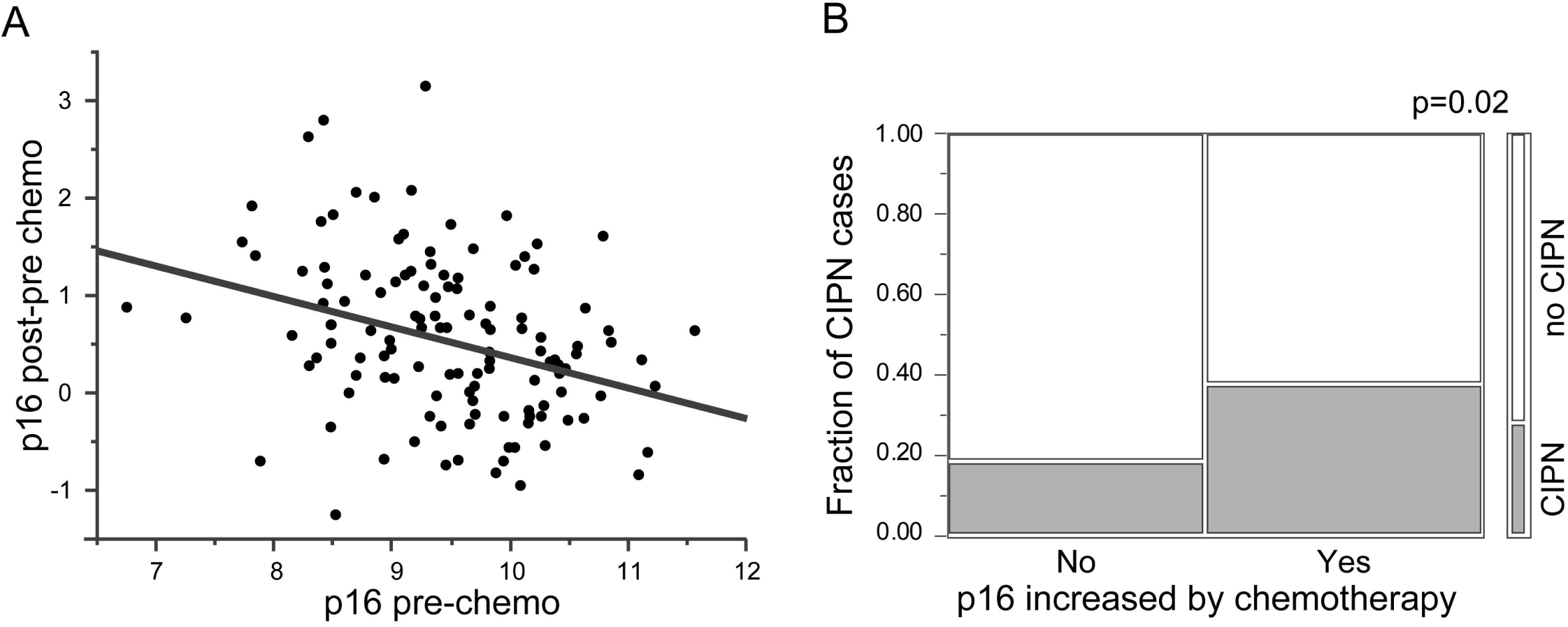
Chemotherapy-induced increase in p16 as risk factor for CIPN. Participants with low p16 were more likely to experience a chemotherapy-induced increase in p16 (A) and were more likely to develop taxane-induced CIPN (B). B. Mosaic plot of incidence of CIPN. CIPN group is shown in red, no CIPN in blue.

## DISCUSSION

Determining which patients are at risk of CIPN is a major unmet need in oncology, given its high incidence and persistence into survivorship. We examined if a measurement of cellular senescence, p16 expression, is a risk factor for CIPN. We anticipated that higher p16 would be associated with greater risk of CIPN as aging could be a risk factor^2^. Surprisingly, we found that patients with the greatest risk were chronologically older but molecularly younger (lower p16 expression). Although our findings of higher CIPN risk for those with age-inappropriately low p16 (low p16Age Gap) initially seemed counterintuitive, this paradoxical result can be understood in the context of data demonstrating that patients with lower baseline p16 expression are more likely to have a larger chemotherapy-induced increase in p16^49^ (Figure 3A). And larger chemotherapy-induced change in p16 expression (pre-treatment to post-treatment) is associated with higher CIPN risk (Figure 3B).

Lower p16Age Gap association with higher risk of CIPN is unlikely to be driven by taxane dose delivered. While total dose was not availbale in this dataset, addition of taxane regimen (weekly vs every 3 weeks) to Model 2 did not change model performance (Supplemental Table S1), and p16Age Gap levels did not differ between participants whose chemotherapy was discontinued or reduced due to toxicities (Supplemental Fig S1). A consideration of total dose delivered is planned for a follow-up study.

Presently, it is unclear why chemotherapy-induced p16 expression is modulated by p16 levels prior to chemotherapy. Tsygankov et al showed that p16 expression plateaus in late middle age after increasing exponentially with age in early to mid-adulthood^50^.

Thus, participants with lower baseline p16 levels may have the capacity for a larger increase in p16 following chemotherapy, while participants with higher baseline p16 levels may have already reached a maximum threshold for senescent cell accumulation, suggesting that p16 levels cannot increase further without causing morbidity. p16Age Gap, measured at baseline, can therefore be interpreted as a predictor of patients that will accumulate more senescent cells in response to chemotherapy, leading to higher p16 expression and CIPN. Regardless of mechanism, our results, once validated, would allow identification of high risk patients, who could then consider CIPN prevention options like chemotherapy regimens without taxanes, or substituting docetaxel for paclitaxel. Such high-risk patients would also be ideal candiates to be included in trials evaluating CIPN preventive strategies.

While our study provides the first evidence of the connection between p16, senescence, and CIPN in humans, a causal relationship between senescence and CIPN has been demonstarted in mice. Removal of the p16+ senescent cells in oxaliplatin-treated mice completely reversed symptoms of CIPN^32^. Although these discoveries may lead to ways to prevent CIPN through depletion of senescent cells (i.e. senolytic therapies), much work remains to understand human senescence and identify safe and efficacious senolytic therapies.

In this study, we did not find diabetes, obesity, or age to be important contributors to multivariate models of CIPN. A larger study is now underway (SENSE, NCT04932031) that will provide the opportunity to validate p16Age Gap findings as well as interrogate variables not available in this study such as taxane dose intensity, severity of co-morbidities, genetics, drug metabolism, and functional performance to a CIPN prognostic model. In addition, there is great interest in genetic polymorhisms as predictors of CIPN with different chemotherapeutic agents, including taxanes, which may ulimately prove of great utility.^51^

In addition to the sample size, another limitation of this study is that measurement of p16Age Gap was calculated using linear regression of p16 versus age but, as noted above, the best fit between p16 and chronological age is likely non-linear later in life. This may explain the extreme negative p16Age Gap values for some patients in our analysis. We are currenlty conducting a study to build a computational model of senescence in various patient cohorts to provide a better estimator of p16Age Gap. But regardless of the absolute value for the p16Age Gap, low p16 expression in patients with higher chronologic age is a significant risk factor.

Our ongoing studies will refine a p16Age Gap-based model for CIPN prediction into a lab-developed test to be used clinically to obtain a CIPN risk score. In early-stage breast cancer patients, this score can help to guide chemotherapy selection, given that regimens for this indication usually employ one of two different taxanes (paclitaxel or docetaxel) with similar efficacy but different risks of CIPN incidence^5,27^. Patients with age-inappropriate low p16 may be offered docetaxel regimens, possibly in combination with other efforts to reduce CIPN such as cryotherapy, or closer monitoring for CIPN symptoms^23–26^. Additionally, anthracycline regimens that lack taxanes but have similar efficacy might be preferable for pateints where even moderate risk of loss of function due to neuropathy may be unacceptable (for example musicians, surgeons, artists).

In summary, measures of senescent cell load could ultimately help guide clinicians to avoid dose-limiting toxicities and improve quality of life in breast cancer patients. These observations may also be clinically relevant in other cancers that are treated with neurotoxic chemotherapies.

## Data Availability

All data produced in the present work are contained in the manuscript

## ACKNOWLEDGMENTS

This work was supported by grants from Breast Cancer Research Foundation (New York, NY), Kay Yow Cancer Fund (Raleigh, NC), UNC Lineberger Comprehensive Cancer Center/University Cancer Research Fund (Chapel Hill, NC) and NIH/NCI R01 CA203023. The funding sources had no involvement in the study design or in the collection, analysis and interpretation of data. The funding sources also had no involvement in the writing of this report or in the decision to submit the report for publication.

## Author Contributions

Drs Muss, Nyrop, Mitin had full access to all of the data in the study and take responsibility for the accuracy of the data analysis.

Concept and design: Muss, Mitin, Nyrop.

Acquisition, analysis, or interpretation of data: Muss, Nyrop, Reinbolt, Speca, Kimmick, Karuturi, Knecht, Strum, Mitin.

Drafting of the manuscript: Mitin, Muss.

Critical revision of the manuscript for important intellectual content: All authors.

Statistical analysis: Mitin.

Obtained funding: Muss.

