## Supplemental Information for "A biomarker of aging, p16, predicts peripheral neuropathy in women receiving adjuvant taxanes for breast cancer"

Table S1. Addition of taxane administration frequency to the p16Age Gap-driven model does not change model performance.

| **Variable** | **Estimate** | **p** |
| --- | --- | --- |
| p16Age Gap | -0.047 | 0.01 |
| p16 pre-chemo | 1.25 | 0.04 |
| Taxane type [paclitaxel] | 0.88 | 0.002 |
| Taxane frequency [weekly] | -0.24 | 0.32 |
| **Model Performance** |  |  |
| AICc | 158 |  |
| BIC | 172 |  |
| Chi-square | 29.5 |  |
| p value | 0.0001 |  |

Table S2. Dose reduction and discontinuation due to toxicities.

|  | **All**  **N=152** | **Grade 2+ CIPN**  **n=44** | **No CIPN**  **n=108** |
| --- | --- | --- | --- |
| Dose reduction, n (%)  Dose discontinuation, n (%) | 46 (30); 22 (14) | 18 (41); 10 (23) | 28 (26); 12 (11) |

Figure S1. P16Age Gap does not differ between patients groups whose taxane regimen was discontinued or reduced due to chemotherapy toxicity.

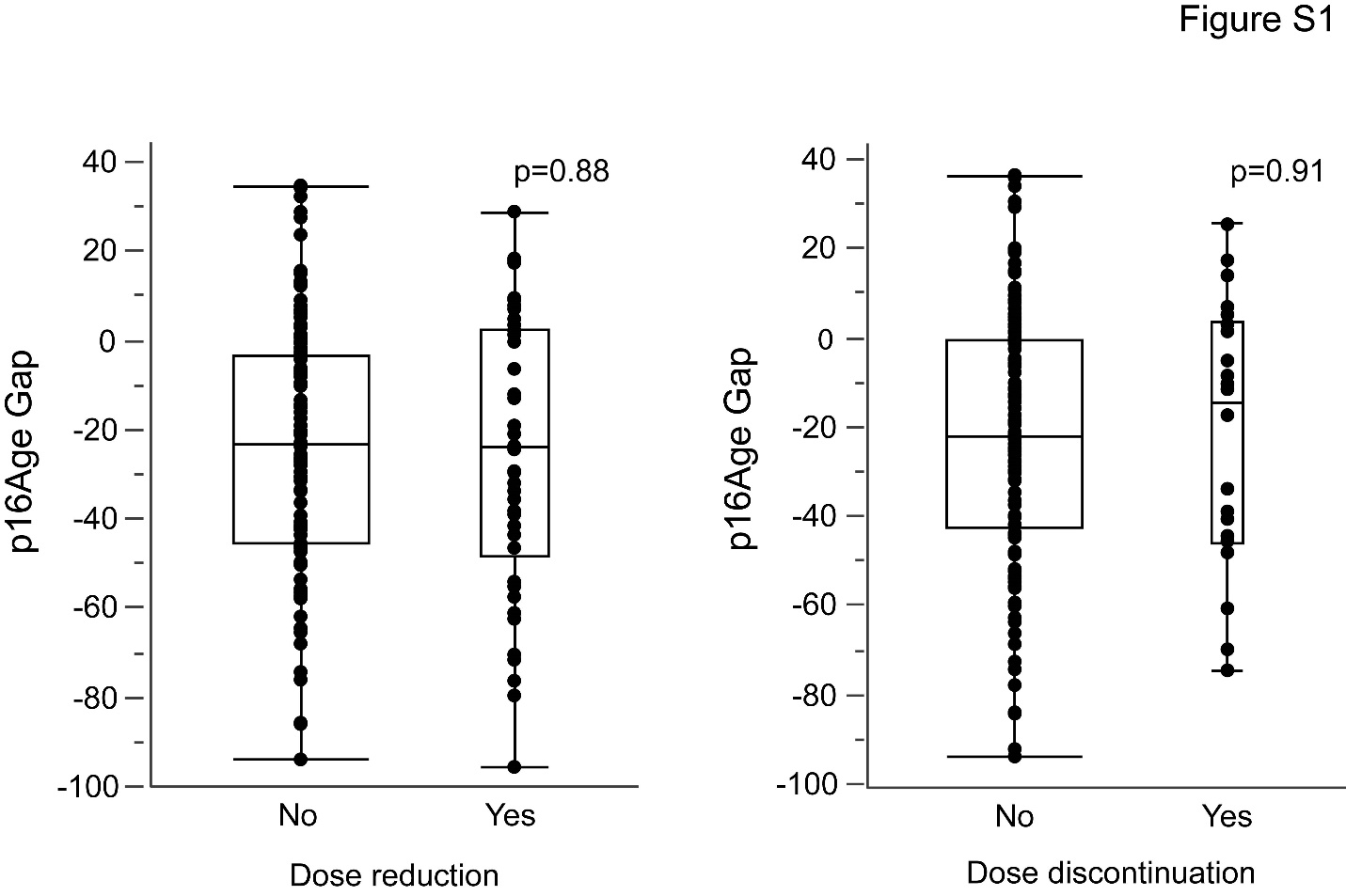
